# Molecular and Immunological Signatures of Long COVID: Implications for Diagnosis and Personalized Treatment Strategies

**DOI:** 10.1101/2025.02.02.24318641

**Authors:** Jorine K.N. Hammink, Tim R. van Elst

## Abstract

The COVID-19 pandemic, caused by the SARS-CoV-2 virus, has led to an emerging health challenge known as “long COVID,” also known as PASC, post-acute sequelae of COVID-19, characterized by symptoms that persist beyond the acute phase of infection. While acute COVID-19 has been extensively studied, the molecular and immunological mechanisms underlying long COVID remain poorly understood. This study aims to investigate these mechanisms by examining the presence of the viral nucleocapsid (N) and spike (S) genes, their mRNA expression, associated immunoglobulins (IgG), and immune regulation via IDO-2 activity in blood of individuals suspected of long COVID. Here we show that a unique pattern of test results contributes to a better understanding of the underlying mechanisms of long COVID, ultimately leading to improved diagnostic and therapeutic strategies for this condition. This study focuses on four key objectives: detecting viral or vaccin induced genetic material, quantifying mRNA expression of the N and S genes, profiling immunoglobulin levels, and measuring IDO-2 activity. These objectives aim to differentiate long COVID from other post-infectious conditions and provide insights into prolonged symptoms. The study population comprised 72 participants, 31 of whom were suspected of having long COVID based on defined symptomatology. Viral genetic material was detected in both symptomatic and asymptomatic individuals, Immunoglobulin levels varied, with symptomatic males exhibiting lower anti-Spike IgG levels than females, suggesting possible gender differences in immune response. Logistic regression models revealed that mRNA spike data alone in this small group was insufficient to predict symptoms presence, but the inclusion of immunoglobulins and inflammatory markers significantly improved predictive accuracy.

Overall, this study highlights the complexity of long COVID and suggests that a multi-variable approach, combining mRNA and genomic spike data with inflammatory markers and demographic factors, provides a basis for effective prediction of symptoms, helping refine diagnostic and therapeutic strategies for long COVID

## Introduction

The COVID-19 pandemic, caused by the SARS-CoV-2 virus, has precipitated an unprecedented global health crisis [1]. While the acute phase of the infection has been extensively studied [2,3], there is growing recognition of “long COVID,” characterized by prolonged symptoms that persist beyond the acute phase of infection [4,5]. These lingering symptoms affect a substantial number of COVID-19 survivors, with evidence suggesting that long COVID could stem from complex molecular and immunological mechanisms that extend beyond active viral replication [6,7].

This study aims to elucidate the molecular and immunological underpinnings of long COVID by investigating the simultaneous presence of the COVID-19 nucleocapsid (N) gene and the spike (S) gene, their respective mRNA expression products, associated immunoglobulins indicative of past infection or vaccination, and the immune system’s resting state through IDO-2 measurements. The SARS-CoV-2 N gene encodes the nucleocapsid protein, essential for viral RNA encapsulation [8], while the S gene encodes the spike protein, which mediates viral entry into host cells via the ACE2 receptor [9,10]. The simultaneous detection of the N and S genes, even in recovered patients, suggests a possible role for persistent viral fragments in the ongoing symptomatic process, providing new insights into long-term viral impact [11].

Our primary hypothesis posits that long COVID represents a distinct clinical syndrome identifiable by unique combinations of test results that cannot be adequately explained by classical infection paradigms. This hypothesis arises from observations that some individuals exhibit prolonged symptoms and immunological markers indicative of an ongoing pathological process [12,13], despite the absence of active viral replication detectable by standard diagnostic methods [14]. There is growing evidence that residual viral particles or immune responses may contribute to long-term symptoms, even after the virus is no longer infectious [15,16].

This study is centered on four primary objectives:

1. Detection of Viral Genetic Material: To detect the presence of the COVID-19 N and S genes in samples from individuals suspected of long COVID. The presence of these genes would indicate residual viral particles or fragments that might be contributing to ongoing symptoms [17].
2. messenger RNA Expression Analysis: To measure the expression levels of mRNA corresponding to the N and S genes. Elevated levels of these mRNAs might suggest persistent viral activity or delayed clearance of viral genetic material [18].
3. Immunoglobulin Profiling: To quantify immunoglobulins (IgG) against the N and S proteins. These immunoglobulins serve as markers of past infection or vaccination and might help differentiate between long COVID and other conditions with similar symptoms [19,20].
4. IDO-2 Activity Measurement: To assess the resting state of the immune system through IDO-2 (indoleamine 2,3-dioxygenase 2) activity. IDO-2 is an enzyme involved in immune regulation, and its activity can provide insights into the immune system’s state, particularly in relation to chronic inflammation and immune exhaustion [21].

These research objectives will collectively contribute to a better understanding of the underlying mechanisms of long COVID, ultimately leading to improved diagnostic and therapeutic strategies for this condition. By detecting viral genetic materials and analyzing mRNA expression, researchers may determine whether there is persistent viral activity or delayed clearance of viral material [22]. Immunoglobulin profiling will assist in identifying past infections or vaccinations and in distinguishing long COVID from other conditions [23]. Finally, measuring IDO-2 activity will provide crucial information on the state of the immune system, which is vital for understanding chronic inflammation and immune exhaustion associated with long COVID [24].

## Methods

### Study population

This study population consisted of 72 persons (35 men and 37 women) in the age of 18–89 year. The test subjects were divided in a potential long COVID group based on long COVID specific symptomatology as described by the National Institute for Public Health and the Environment, Bilthoven, The Netherlands (fatigue, shortness of breath, chest pain, muscle aches, headaches, heart palpitations, prolonged loss of smell, depression or memory problems, https://www.rivm.nl/en/coronavirus-covid-19/long-covid) and a control group. Test subjects which did not present this particular symptomatology were assigned to the control group The potential long COVID group consisted of 31 subjects and the control group consisted of 41 subjects.

### Ethics statement

All subjects included used the commercial services of NL-Lab, Leeuwarden, The Netherlands to detect the presence of SARS-CoV-2 in their blood and signed and approved for the use of their archived and fully anonymized saliva samples for research purposes. Since the blood samples were anonymized diagnostic surplus material, no approval from the Medical Research Ethics Committee was needed, according to the Code of conduct for responsible use (Federation of Dutch Medical Scientific Societies).

### Sample Collection and Preparation

Blood samples were collected from participants diagnosed with long COVID and control individuals. Blood samples were drawn into EDTA tubes to prevent coagulation. All samples were processed within two hours to maintain the integrity of nucleic acids and proteins.

### Computerized Fluorescence in situ Hybridization (C-FISH)

The detection and quantification of SARS-CoV-2 nucleocapsid (N) and spike (S) genes were performed using the Computerized-Fluorescent In Situ Hybridization (C-FISH) technique (NL-Lab, The Netherlands). Specific probes complementary to the N and S gene sequences were designed based on the latest sequence data from the GISAID database. The C-FISH protocol involved hybridizing these fluorescently labeled probes to the target sequences in the samples, followed by visualization and quantification using a fluorescence microscope and image analysis software [11].

### mRNA Expression Analysis

The expression levels of N and S gene mRNAs were quantified using C-FISH (NL-Lab, The Netherlands) with the same but complementary DNA probes used for gene hybridization. Fluorescence intensity corresponding to the hybridized probes was measured, providing a quantitative assessment of mRNA expression levels.

### Immunoglobulin Quantification

Levels of specific immunoglobulins (IgG, IgA, and IgM) against the N and S proteins were quantified using enzyme-linked immunosorbent assay (ELISA). Blood plasma was separated by centrifugation at 1500 g for 10 minutes and stored at −80°C until analysis. ELISA plates were coated with recombinant N and S proteins (Sino Biological, China) and incubated with plasma samples. Bound immunoglobulins were detected using horseradish peroxidase (HRP)-conjugated secondary antibodies and TMB substrate solution. Absorbance was measured at 450 nm using a microplate reader (Bio-Rad, USA), and concentrations were determined using standard curves generated with known quantities of immunoglobulins.

### IDO-2 Activity Assay

IDO-2 activity was measured using a commercially available kynurenine assay to quantify the conversion of tryptophan to kynurenine, which serves as an indicator of immune activation and regulatory status. Plasma samples were deproteinized with 10% trichloroacetic acid and incubated with Ehrlich’s reagent. The resulting colorimetric change, corresponding to kynurenine concentration, was measured at 492 nm using a microplate reader (Bio-Rad, USA). A standard curve of kynurenine was used to calculate the IDO-2 activity in the samples. This assay provides insights into the immune system’s regulatory mechanisms, particularly in relation to chronic inflammation and immune exhaustion.

### Statistical analysis

A multi-parameter logistic regression model was constructed to predict a binary outcome (symptom presence or absence) using multiple independent variables, including mRNA spike levels, genomic IgG markers (S-IgG, N-IgG), an inflammatory biomarker (IDO-2), and demographic factors (age, gender, vaccination status). Maximum likelihood estimation (MLE) was employed to estimate the coefficients of the independent variables. The model assumes a linear relationship between the log-odds of the outcome and the predictors. Predictor multi-collinearity was assessed, and the model’s goodness-of-fit was evaluated using metrics such as the Akaike Information Criterion (AIC) and likelihood ratio tests.

## Results

### Description of dataset

This study analyzed 72 participants, consisting of 35 males and 37 females. Among the males, 83% were symptomatic, compared to 51% of females, indicating a higher proportion of symptomatic males. table 2 shows patient characteristics. Although the genomic nucleocapsid (N) detection was infrequent, the presence of N in both symptomatic and asymptomatic participants suggests that viral particles or remnants may still be detectable, which could relate to the persistence of some symptoms. Nucleocapsid (N) detection was found in only 1 symptomatic male and 6 asymptomatic males, while 4 symptomatic and 4 asymptomatic females were positive. Similar trends were observed for mRNA N detection, with asymptomatic participants showing higher positivity. However, the spike (S) gene detection, especially in symptomatic individuals, hints at a lingering viral presence that could contribute to the ongoing symptomatology. Anti-Nucleocapsid IgG was not detected in any participant, but anti-Spike IgG (S IgG) levels varied, with symptomatic males having lower levels (mean = 10) compared to females (mean = 15), suggesting gender differences in immune response. Levels of IDO-2 were consistently present across all groups, with no significant differences between symptomatic and asymptomatic participants. See table 2.

**Table 1.**
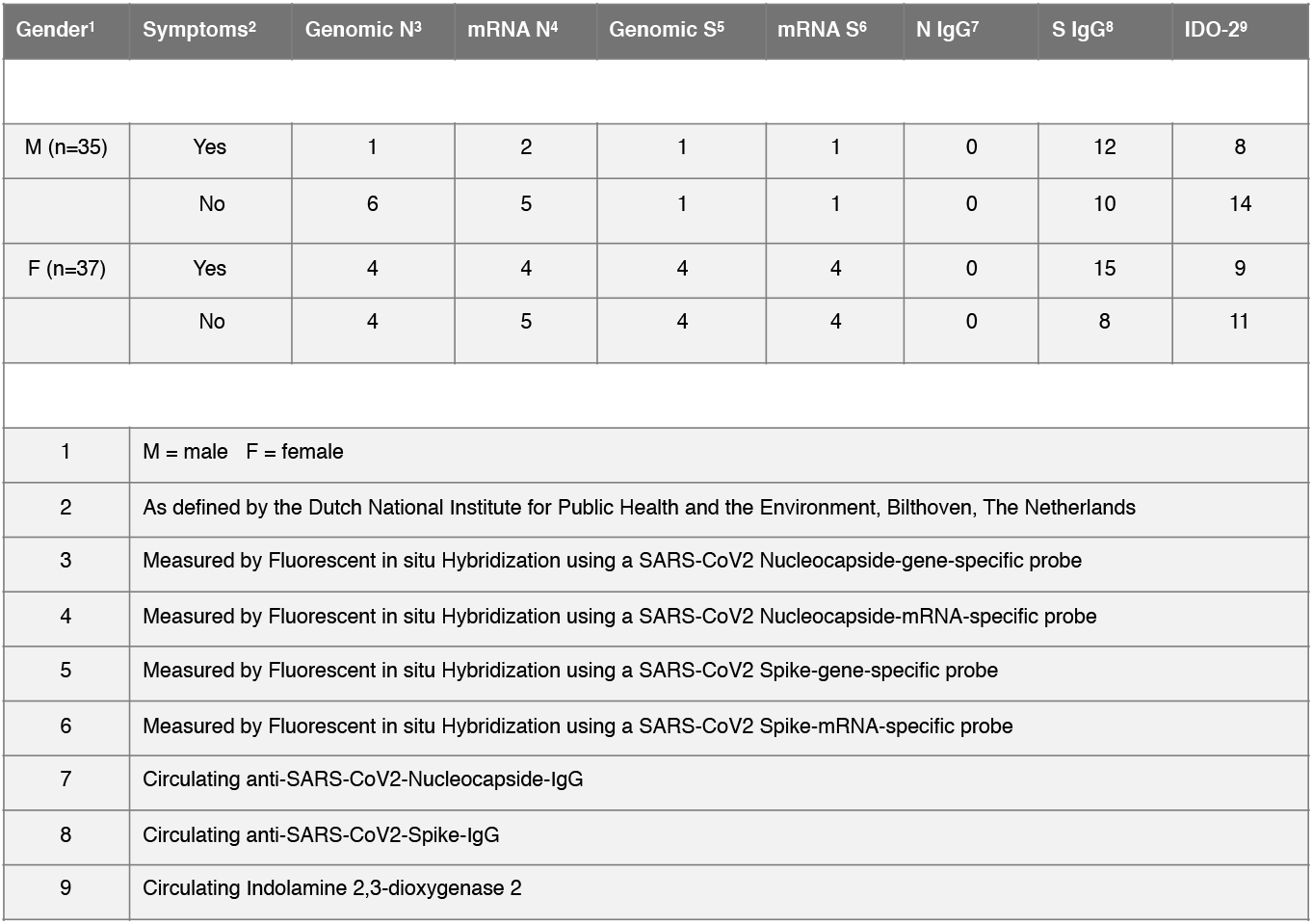
In the frequency distribution the data obtained during this study is listed.

| Gender<sup>1</sup> | Symptoms<sup>2</sup> | Genomic N<sup>3</sup> | mRNA N<sup>4</sup> | Genomic S<sup>5</sup> | mRNA S<sup>6</sup> | N IgG<sup>7</sup> | S IgG<sup>8</sup> | IDO-2<sup>9</sup> |
| --- | --- | --- | --- | --- | --- | --- | --- | --- |
| M (n=35) | Yes | 1 | 2 | 1 | 1 | 0 | 12 | 8 |
|  | No | 6 | 5 | 1 | 1 | 0 | 10 | 14 |
| F (n=37) | Yes | 4 | 4 | 4 | 4 | 0 | 15 | 9 |
|  | No | 4 | 5 | 4 | 4 | 0 | 8 | 11 |
| 1 | M = male F = female |  |  |  |  |  |  |  |
| 2 | As defined by the Dutch National Institute for Public Health and the Environment, Bilthoven, The Netherlands |  |  |  |  |  |  |  |
| 3 | Measured by Fluorescent in situ Hybridization using a SARS-CoV2 Nucleocapside-gene-specific probe |  |  |  |  |  |  |  |
| 4 | Measured by Fluorescent in situ Hybridization using a SARS-CoV2 Nucleocapside-mRNA-specific probe |  |  |  |  |  |  |  |
| 5 | Measured by Fluorescent in situ Hybridization using a SARS-CoV2 Spike-gene-specific probe |  |  |  |  |  |  |  |
| 6 | Measured by Fluorescent in situ Hybridization using a SARS-CoV2 Spike-mRNA-specific probe |  |  |  |  |  |  |  |
| 7 | Circulating anti-SARS-CoV2-Nucleocapside-IgG |  |  |  |  |  |  |  |
| 8 | Circulating anti-SARS-CoV2-Spike-IgG |  |  |  |  |  |  |  |
| 9 | Circulating Indolamine 2,3-dioxygenase 2 |  |  |  |  |  |  |  |

**Table 2.**
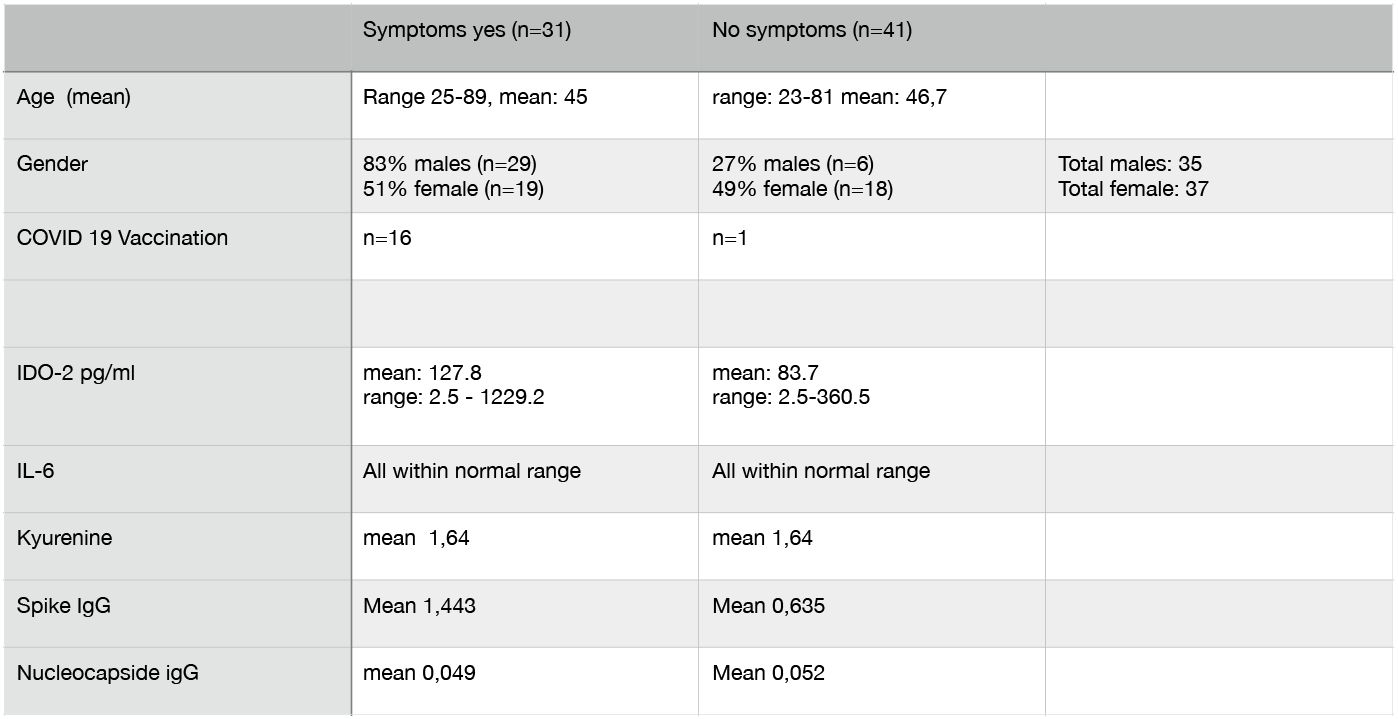
Demographic data of study participants and inflammatory markers.

### Description of logistic regression model

The initial logistic regression model used mRNA spike levels alone as predictors. This model produced an overall accuracy of 52%, with a high recall for the absence of symptoms (recall = 1.00) but no predictive power for the presence of symptoms (precision = 0.00). These results suggest that mRNA spike data, by itself, is insufficient to effectively distinguish between symptomatic and asymptomatic individuals. To improve predictive performance, the model was expanded to include genomic spike data (S-IgG and N-IgG) in addition to the mRNA spike levels. This extended model resulted in an overall accuracy of 65%, with balanced performance in predicting both the absence (precision = 0.67, recall = 0.67) and presence of symptoms (precision = 0.64, recall = 0.64). The inclusion of IgG markers enhanced the model’s ability to predict symptom presence, indicating that immune response markers, such as IgG, are valuable in distinguishing symptomatic individuals. Further refinement of the model was achieved by incorporating additional variables, including inflammatory biomarkers (IL-6, Kynurenine, IDO-2), demographic factors (age, gender), and vaccination status. This comprehensive model yielded an accuracy of 78%, significantly improving the model’s predictive power. In particular, the model was highly effective at predicting the absence of symptoms (precision = 0.73, recall = 0.92). The ability to predict symptom presence also improved, with precision reaching 0.88 and recall at 0.64. The addition of biomarkers and demographic information contributed to a more nuanced understanding of symptom manifestation, leading to greater model accuracy overall.

These results demonstrate that a multi-variable approach, combining mRNA and genomic spike data with inflammatory markers and demographic factors, provides a more effective prediction of symptom presence than using mRNA spike data alone.

## Discussion

This study aimed to investigate the predictive value of laboratory patterns in relation to long COVID-associated symptoms, focusing on viral genetic markers, immune responses, and inflammatory biomarkers. Our findings highlight the complexities of predicting symptomatic manifestations of COVID-19, especially in individuals with long COVID, based on viral detection and immune response markers. The detection of viral genetic material, although not universally associated with symptomatic presentation, points towards a nuanced role of these fragments in the persistence of symptoms. Despite a higher proportion of symptomatic males (83%) compared to females (51%), there was no strong correlation between the presence of genomic nucleocapsid (N) or spike (S) signals and symptom manifestation. However, the appearance of these signals in both symptomatic and asymptomatic individuals suggests that persistent viral material could play a role in symptom development, even in the absence of active viral replication. For example, 5 asymptomatic males had detectable mRNA N signals compared to only 2 symptomatic males, and similar patterns were observed for genomic and mRNA S signals. These results suggest that viral RNA detection, while not conclusive for symptoms alone, may still influence the broader immunological landscape, contributing to the long-term condition. This finding also aligns with emerging evidence suggesting that asymptomatic carriers of SARS-CoV-2 can harbor viral genetic material without developing symptoms, possibly due to differences in host immune response or viral replication dynamics [11]. The comparable detection rates of genomic and mRNA S signals between symptomatic and asymptomatic groups further support this hypothesis. These observations challenge the assumption that viral load or the mere presence of viral genetic material is a reliable predictor of symptomatology, emphasizing the need to explore other biological factors involved in symptom manifestation. The analysis of IgG antibody levels provided additional insights into the role of humoral immunity in symptom development. While anti-Nucleocapsid IgG (N IgG) was undetectable in all participants, anti-Spike IgG (S IgG) levels varied significantly between symptomatic and asymptomatic individuals. See table 2. Notably, symptomatic males had lower mean S IgG levels (mean = 10) compared to symptomatic females (mean = 15). This gender-based disparity suggests that females may mount a stronger humoral immune response against the Spike protein, potentially offering greater protection or mitigating the severity of symptoms. Such differences in immune response could be influenced by hormonal, genetic, or environmental factors that require further investigation. Interestingly, among asymptomatic participants, both males and females exhibited similar S IgG levels, suggesting that while S IgG may play a role in modulating symptoms, it is not the sole determinant of symptomatic presentation. In terms of predictive modeling, the initial logistic regression model that relied exclusively on mRNA spike levels was inadequate in predicting the presence of symptoms, with an accuracy of only 52%. The model performed well in identifying individuals without symptoms (recall = 1.00) but failed to predict symptomatic cases (precision = 0.00). This finding reinforces the notion that mRNA spike levels, in isolation, are insufficient to distinguish between symptomatic and asymptomatic cases, likely due to the presence of asymptomatic carriers with detectable viral RNA. When genomic spike data (S IgG and N IgG) were incorporated into the model, the predictive accuracy improved to 65%, with a more balanced performance in predicting both the presence and absence of symptoms (precision and recall = 0.64 for symptomatic cases, precision and recall = 0.67 for asymptomatic cases). The inclusion of IgG markers suggests that immune responses, particularly those related to the Spike protein, are more predictive of symptomatic manifestation than viral RNA alone. This aligns with the hypothesis that immune responses, rather than viral load or presence, may be key drivers of symptom variability in COVID-19 and long COVID. The most comprehensive model, which integrated an inflammatory biomarker (IDO-2), demographic factors (age, gender), and vaccination status, achieved the highest predictive accuracy of 78%. The model’s ability to predict the absence of symptoms (precision = 0.73, recall = 0.92) was particularly strong, while its performance in predicting symptom presence (precision = 0.88, recall = 0.64) also showed significant improvement. These results highlight the importance of a multi-dimensional approach to symptom prediction, incorporating a wide range of biological markers and demographic factors. Interestingly, levels of circulating IDO-2, an enzyme involved in immune regulation, were consistently present across all participants, with no significant differences between symptomatic and asymptomatic groups. This suggests that IDO-2 may not be a major driver of symptom variability in this cohort, though its role in immune modulation warrants further investigation in larger and more diverse populations. The gender-based differences in immune response observed in this study are also noteworthy. Symptomatic females exhibited higher levels of S IgG compared to symptomatic males, which may suggest a more robust humoral immune response in females. This could partially explain the lower proportion of symptomatic females in the cohort. Gender differences in immune responses have been observed in other infectious diseases and are thought to be influenced by both hormonal and genetic factors. These findings raise important questions about the role of gender in COVID-19 pathogenesis and immune responses, particularly in the context of long COVID.

In conclusion, this study underscores the complexity of predicting COVID-19 symptoms based solely on viral detection. While mRNA and genomic spike signals provide some insight into viral presence (especially in the acute COVID scenario) immune response markers, particularly S IgG, offer greater predictive value for symptom manifestation in case of long COVID. The integration of inflammatory biomarkers, demographic data, and immune responses significantly enhances the accuracy of predictive models, providing a more comprehensive understanding of the factors contributing to symptom variability in COVID-19 and long COVID. Moreover individual outcomes can contribute to personalized treatment strategies in long COVID. Treatment options should be further evaluated in future studies.

## Data Availability

All data produced in the present study are available upon reasonable request to the authors

