## supplemental table 1 for "Molecular and Immunological Signatures of Long COVID: Implications for Diagnosis and Personalized Treatment Strategies"

In Table 1  the frequency distribution the data obtained during this study is listed.

| Gender[1] | Symptoms[2] | Genomic N[3] | mRNA N[4] | Genomic S[5] | mRNA S[6] | N IgG[7] | S IgG[8] | IDO-2[9] |
|---|---|---|---|---|---|---|---|---|
| M (n=35) | Yes | 1 | 2 | 1 | 1 | 0 | 12 | 8 |
| | No | 6 | 5 | 1 | 1 | 0 | 10 | 14 |
| F (n=37) | Yes | 4 | 4 | 4 | 4 | 0 | 15 | 9 |
| | No | 4 | 5 | 4 | 4 | 0 | 8 | 11 |
| 1 | M = male F = female |  |  |  |  |  |  |  |
| 2 | As defined by the Dutch National Institute for Public Health and the Environment, Bilthoven, The Netherlands | | | | | | | |
| 3 | Measured by Fluorescent in situ Hybridization using a SARS-CoV2 Nucleocapside-gene-specific probe | | | | | | | |
| 4 | Measured by Fluorescent in situ Hybridization using a SARS-CoV2 Nucleocapside-mRNA-specific probe | | | | | | | |
| 5 | Measured by Fluorescent in situ Hybridization using a SARS-CoV2 Spike-gene-specific probe | | | | | | | |
| 6 | Measured by Fluorescent in situ Hybridization using a SARS-CoV2 Spike-mRNA-specific probe | | | | | | | |
| 7 | Circulating anti-SARS-CoV2-Nucleocapside-IgG | | | | | | | |
| 8 | Circulating anti-SARS-CoV2-Spike-IgG | | | | | | | |
| 9 | Circulating Indolamine 2,3-dioxygenase 2 | | | | | | | |
