## supplemental table 2 for "Molecular and Immunological Signatures of Long COVID: Implications for Diagnosis and Personalized Treatment Strategies"

Table 2. Demographic data of study participants and inflammatory markers

| | Symptoms yes (n=31) | No symptoms (n=41) | |
|---|---|---|---|
| Age (mean) | Range 25-89, mean: 45 | range: 23-81 mean: 46,7 | |
| Gender | 83% males (n=29)<br>51% female (n=19) | 27% males (n=6)<br>49% female (n=18) | Total males: 35<br>Total female: 37 |
| COVID 19 Vaccination | n=16 | n=1 | |
| IDO-2 pg/ml | mean: 127.8<br>range: 2.5 - 1229.2 | mean: 83.7<br>range: 2.5-360.5 | |
| IL-6 | All within normal range | All within normal range | |
| Kyurenine | mean 1,64 | mean 1,64 | |
| Spike IgG | Mean 1,443 | Mean 0,635 | |
| Nucleocapside igG | mean 0,049 | Mean 0,052 | |
